# Factors Associated with COVID-19 Testing among People who Inject Drugs: Missed Opportunities for Reaching those Most at Risk

**DOI:** 10.1101/2022.01.04.22268749

**Authors:** Samantha Yeager, Daniela Abramovitz, Alicia Harvey-Vera, Carlos F. Vera, Angel B. Algarin, Laramie R. Smith, Gudelia Rangel, Irina Artamonova, Thomas L. Patterson, Angela R. Bazzi, Emma L. Brugman, Steffanie A. Strathdee

**Author notes:** Corresponding Author: Steffanie Strathdee, PhD., Distinguished Professor, Harold Simon Chair, Associate Dean of Global Health Sciences. Division of Infectious Diseases and Global Public Health, Department of Medicine, University of California San Diego, 9500 Gilman Drive, Mail Code 0507, La Jolla, CA 92093-0507.

## Abstract

People who inject drugs (PWID) are vulnerable to SARS-CoV-2 infection. We examined correlates of COVID-19 testing among PWID in the U.S.-Mexico border region and described encounters with services or venues representing potential opportunities (i.e., ‘touchpoints’) where COVID-19 testing could have been offered. Between October, 2020 and September, 2021, participants aged ≥18 years from San Diego, California, USA and Tijuana, Baja California, Mexico who injected drugs within the last month completed surveys and SARS-CoV-2, HIV, and HCV serologic testing. Logistic regression was used to identify factors associated with COVID-19 testing prior to enrollment. Of 583 PWID, 30.5% previously had a COVID-19 test. Of 172 PWID who tested SARS-CoV-2 seropositive in our study (30.1%), 50.3% encountered at least one touchpoint within the prior six months where COVID-19 testing could have been offered. Factors independently associated with at least two fold higher odds of COVID-19 testing were living in San Diego (versus Tijuana), having recently been incarcerated or attending substance use disorder (SUD) treatment and having at least one chronic health condition. In addition, recent homelessness, having had at least one COVID-19 vaccine dose and having been tested for HIV or HCV since the pandemic began were independently associated with COVID-19 testing. We identified several factors independently associated with COVID-19 testing and multiple touchpoints where COVID-19 testing could be scaled up for PWID, such as SUD treatment programs and syringe service programs. Integrated health services are needed to improve access to rapid, free COVID-19 testing in this vulnerable population.

## Introduction

Testing for SARS-CoV-2 infection is critical to identify cases who require quarantine and contact tracing, as well as treatment and supportive housing. Within the United States (U.S.), COVID-19 testing based on polymerase chain reaction (PCR) has been made available at community clinics, pharmacies and laboratories since early in the pandemic.[1] However, PCR tests are expensive and turn-around times for results can take days. The nation’s first rapid at-home COVID-19 testing kit received emergency-use authorization in October, 2020, [2] but rapid tests are not yet widely available and are seldom free. In Mexico, free COVID-19 PCR tests are available at designated testing facilities or “fever clinics” for qualifying individuals, and at private hospitals, laboratories or clinics for a fee. [3, 4] Despite efforts to increase the accessibility of COVID-19 testing, utilization remains low among racial/ethnic minorities and economically disadvantaged populations due to barriers related to health insurance, availability of testing sites, language, transportation, and misinformation. [5–9]

COVID-19 misinformation is a prominent barrier to testing among African American and Latinx communities. [6, 10] Research within the U.S. and United Kingdom has also highlighted connections between COVID-19 disinformation (e.g., conspiracy theories) and lower engagement in preventive behaviors (e.g., handwashing, mask wearing, and social distancing) and vaccination. [11, 12] In a previous analysis, COVID-19 disinformation was significantly associated with COVID-19 vaccine hesitancy among people who inject drugs (PWID) in the U.S.-Mexico border region. [12] However, it remains unknown if COVID-19 misinformation or disinformation impacts COVID-19 testing utilization among PWID.

Due to their high prevalence of chronic diseases, [13] homelessness, [14, 15] food insecurity, [16] frequent incarceration, [17, 18] and behavioral risk factors [(e.g., engaging in sex work, sharing needles with others, [17] PWID are at elevated risk for SARS-CoV-2 infection and developing severe symptoms. [13, 17, 19] PWID often underutilize healthcare services due to stigma and mistreatment [14, 20, 21] which may have been exacerbated during the COVID-19 pandemic due to anticipated stigma (i.e., expectation of future stigma experiences) or enacted stigma (i.e., experiences of stigmatization). [22] However, PWID could receive COVID-19 testing through intersecting venues or touchpoints including substance use disorder (SUD) treatment programs, syringe service programs (SSPs), emergency rooms, and jail/prisons. [23–26]

We identified correlates of COVID-19 testing and described interactions with services or venues where COVID-19 testing could have been offered, drawing from literature on overdose prevention ‘touchpoints’ within the healthcare system.[27] We hypothesized that socio-structural determinants (e.g., food/housing insecurity, Latinx ethnicity), COVID-19 disinformation, and COVID-related stigma (anticipated and enacted stigma) would be associated with less COVID-19 testing. We also hypothesized that PWID with chronic health conditions and those who had recently been incarcerated or received health care services would be more likely to have had a COVID-19 test.

## Methods

### Participants and Eligibility

Between October 28, 2020 and September 10, 2021, adults aged ≥18 or older who injected drugs within the last month and lived in San Diego County or Tijuana were recruited through street outreach, as previously described. [17] Protocols were approved by institutional review boards at the University of California San Diego and Xochicalco University in Tijuana. The study was carried out in accordance with The Code of Ethics of the World Medical Association (Declaration of Helsinki) for research involving humans. Informed consent was obtained from all human participants.

### Survey Measures

After providing written informed consent, participants underwent face-to-face interviewer-administered surveys at baseline and approximately one week later using computer assisted personal interviews. Surveys assessed socio-demographics, injection and non-injection use of specific drugs (ever and in the last six months), chronic health conditions (e.g., diabetes, asthma, hypertension), generalized anxiety disorder symptoms (through the GAD-7), [28] food insecurity, [29] COVID-19 experiences, exposures, and protective behaviors (e.g., social distancing, masking). Perceived threat of COVID-19 was assessed by asking participants how worried they were about getting COVID-19 (or getting it again) on a ten point scale. [30] Participants were compensated $20 USD.

We asked participants if they had ever received a COVID-19 test, and if so, to specify the date, location where the test was conducted and the result (if known). We also inquired about encounters with potential COVID-19 testing touchpoints (i.e., where COVID-19 testing could have been offered) in the last six months. [27] These included being enrolled in a SUD treatment program, having been incarcerated, sleeping in a shelter, using a SSP, having an overdose, or having been tested for HIV or HCV since the COVID-19 epidemic began.

To assess COVID-19 misinformation, we presented participants with seven statements about SARS-CoV-2 transmission, severity, immunity, symptoms, treatments and vaccines and asked them to classify each statement as “True”, “False,” or “Unsure”. These included “COVID-19 is about as dangerous as having the flu” and “many thousands of people have not died from COVID-19.” We then created a binary variable for each statement indicating whether the participant was misinformed or not.

COVID-19 disinformation was assessed through a scale including six conspiracy theory items as previously described. [12] These included “COVID-19 was created by the pharmaceutical industry” or “the Chinese government”, “childhood vaccines cause autism”, [31] as well as three additional items: “COVID-19 vaccines include a tracking device”, “alter DNA”, and “COVID-19 vaccines offered to ‘people like me’ are not as safe”. We dichotomized responses to indicate endorsement of disinformation (“True” and “Unsure”) or not (“False”) and summed them into a total score ranging from 0 to 6.

We assessed COVID-19 related stigma utilizing four key questions adapted from the Every Day Discrimination Scale. [32] Two questions assessed whether participants were worried about experiencing increased verbal/physical harassment during the COVID-19 pandemic because there were fewer people in public or for wearing a mask (i.e., anticipated stigma). The average of these two questions were combined to create the COVID-19 harassment/worry stigma subscale. Two questions assessed potential experiences of increased verbal/physical harassment during the COVID-19 pandemic due to fewer people in public or wearing a mask (i.e., enacted stigma). The average of these two questions were utilized to create the COVID-19 harassment experiences subscale.

### SARS-CoV-2 Antibody Detection

Blood samples were collected by venipuncture. Serum samples were batched and tested weekly by Genalyte® (San Diego, CA), using their Maverick™ Multi-Antigen Serology Panel [33] that detects IgG and IgM antibodies to five SARS-CoV-2 antigens. A machine learning algorithm was used to call results using the Random Forest Ensemble method with 3000 decision trees.[34]

### HIV and HCV Serology

Rapid HIV and HCV tests were conducted using the Miriad^®^ HIV/HCV Antibody InTec Rapid Anti-HCV Test (Avantor, Radnor, PA). Reactive and indeterminate tests underwent a second rapid test with Oraquick^®^ HIV or Oraquick^®^ HCV, respectively (Orasure, Bethlehem, PA) and were confirmed by Western Blot at the UC San Diego Centers for AIDS Research.

### Statistical Analysis

The outcome for this analysis was reporting whether or not they had received a COVID-19 test prior to joining the study (yes/no). Characteristics of participants who had a COVID-19 test versus those who had not were compared using Mann-Whitney U tests for continuous variables and Chi-square tests or Fisher’s exact tests for categorical variables. Univariate and multivariable logistic regressions with robust standard error estimation via generalized estimating equations were performed to identify factors associated with COVID-19 testing.

Variables attaining significance at α=0.10 in univariate regression models were considered for inclusion in multivariable models, using Hosmer and Lemeshow’s purposeful selection of variables approach [35] to arrive at a final model. Variables were retained based on statistical significance and relationships among potential predictors (e.g., correlations, confounding, and interactions). Since availability of COVID-19 testing may have changed during the 11 month study period, we included a linear term representing the time that had elapsed since the interview. Since there was no interaction by site (i.e., residence in San Diego versus Tijuana), we did not stratify by place of residence. All statistical analyses were conducted using SAS, version 9.4.

## Results

### Sample Characteristics and COVID-19 Testing History

The 583 participants who completed baseline and supplemental interviews and responded to questions about COVID-19 testing history were included in this analysis. The majority identified as male (74.3%) and Hispanic, Latinx, or Mexican (73.6%) and 34% had completed high school or its equivalent. By design, approximately half (58.7%) resided in San Diego County (Table 1). Mean age was 43 years (standard deviation [SD]: 11).

**Table 1.**
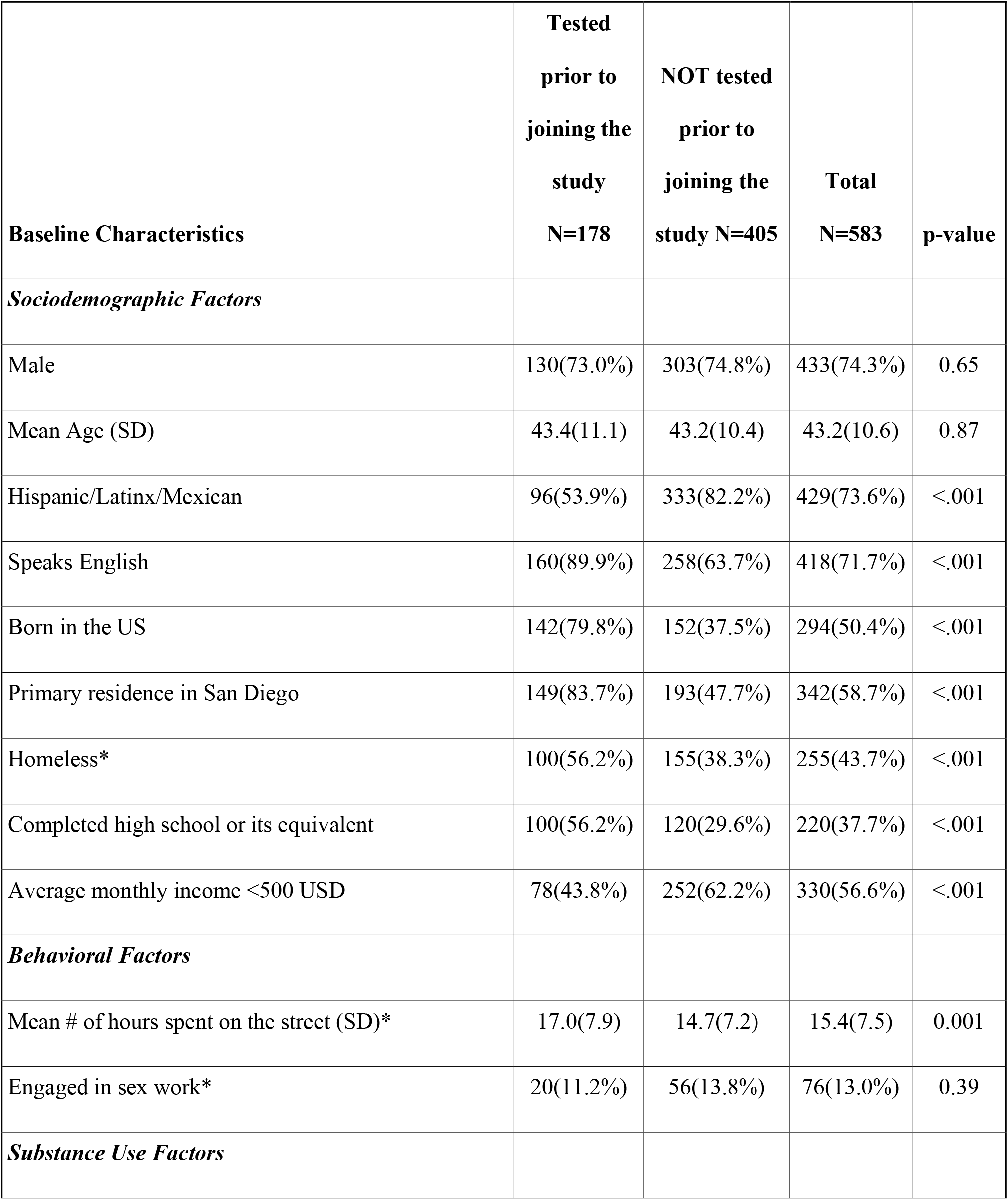
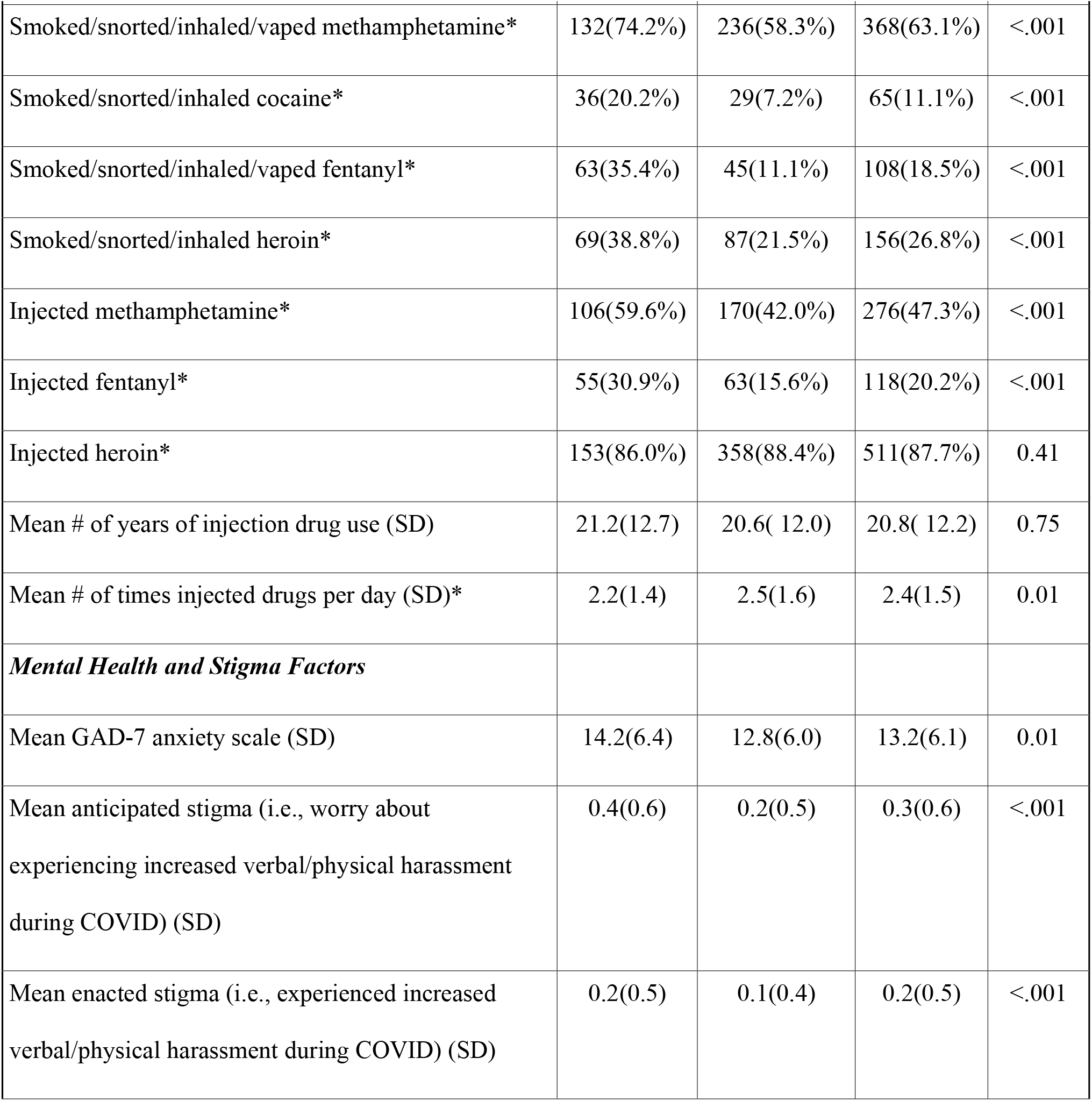
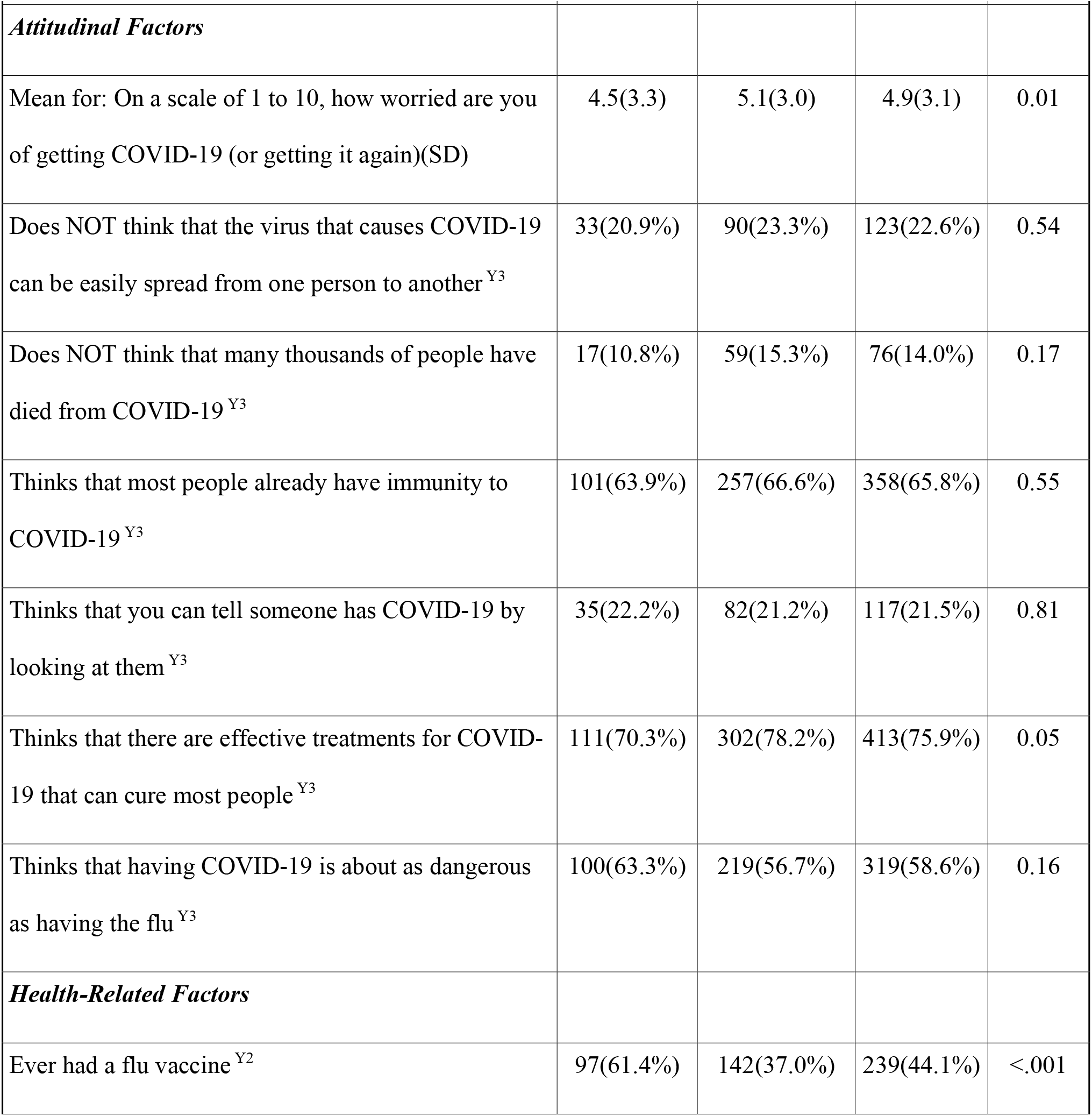
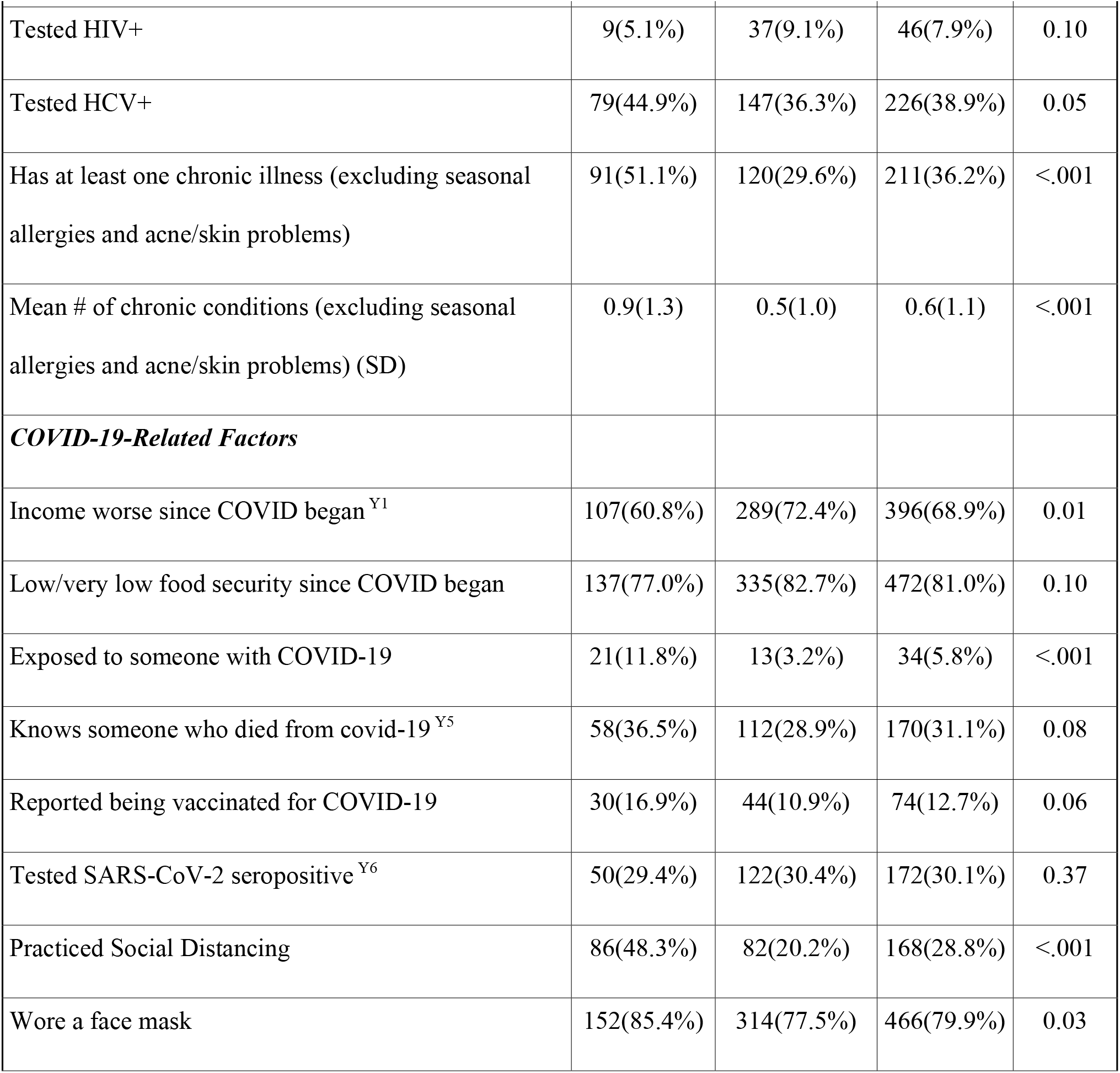
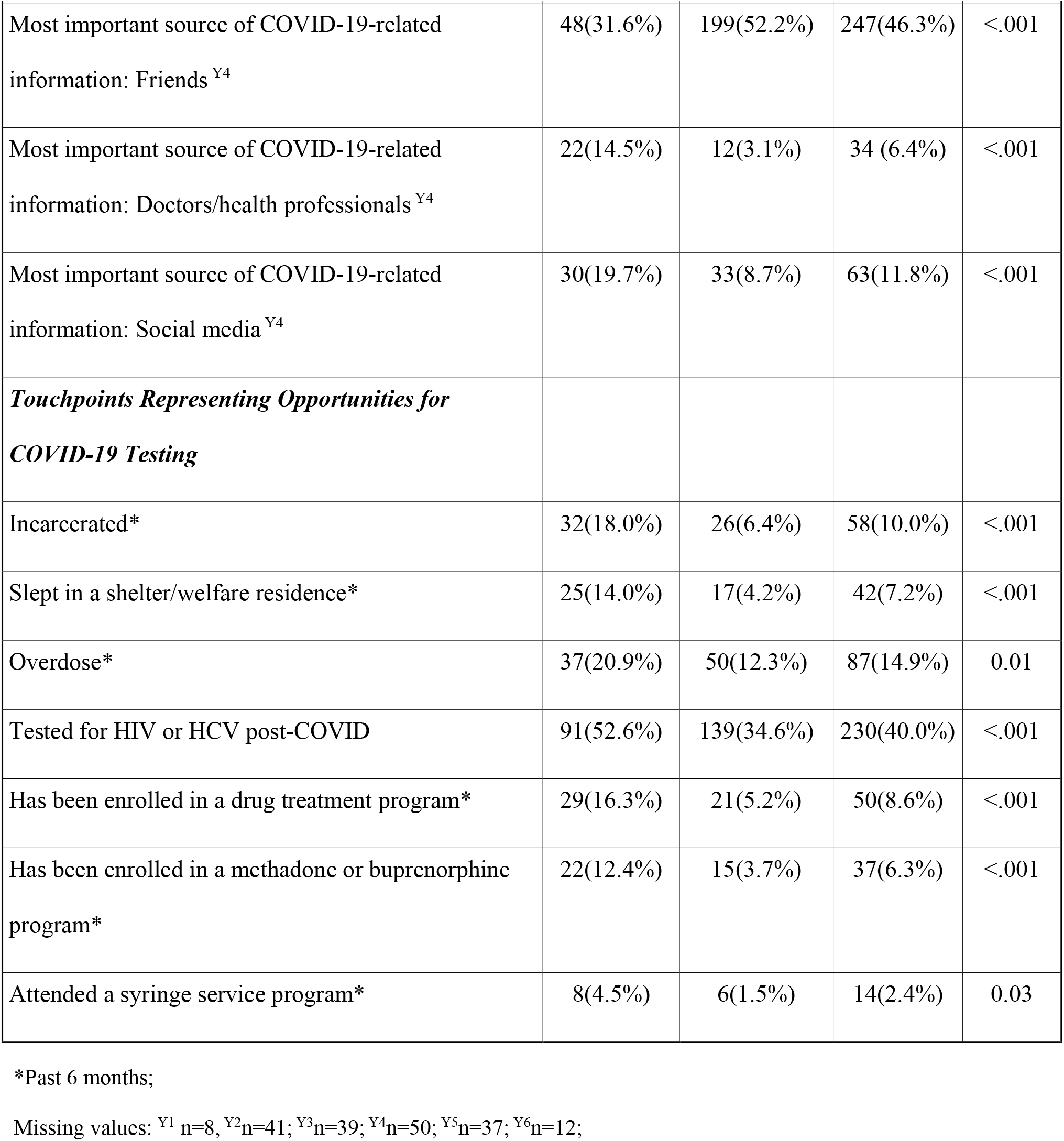
Characteristics Associated with COVID-19 Testing among PWID in San Diego, CA and Tijuana, Mexico (N=583)

In the past six months, 44% of participants were homeless. The majority injected heroin (87.7%), methamphetamine (47.3%) or fentanyl (20.2%) in the last six months. Most had also smoked, snorted or inhaled or methamphetamine (63%), heroin (27%), fentanyl (19%) or cocaine (11%) in the last six months. Over one third tested HCV-seropositive (39%), 8% tested HIV-seropositive and 36% reported at least one other chronic health condition (e.g., diabetes, hypertension).

Overall, 178 participants (30.5%) reported that they previously had a COVID-19 test. Of 105 participants who were asked the location of their COVID-19 test in a supplemental survey, the most common testing locations were community clinics (including mobile clinics and health fairs; 55.2%), hospitals (14.3%), doctors’ offices (14.3%), jail/prison/detention centers (10.5%), SSPs (5.7%), SUD treatment clinics (2.9%), and pharmacies (1.9%).

Considering potential touchpoints for COVID-19 testing, 40% had received an HIV or HCV test outside of the study since the COVID-19 epidemic began. In the last 6 months, 15% had an overdose, 10% had been incarcerated, 7.2% slept in a shelter or a welfare residence, 8.6% had visited a SUD treatment program and 2.4% had used a SSP. Of the 405 participants who had not had a prior COVID-19 test, almost half (46%) reported encounters with at least one touchpoint where COVID-19 testing could have been offered. Furthermore, of 571 participants who provided blood samples for SARS-CoV-2 serology and who tested seropositive in our study (N=172, 30.1%), 70.9% had not previously had a COVID-19 test and 50.3% had encounters with at least one touchpoint where COVID-19 testing could have been offered.

### Factors associated with COVID-19 Testing in Bivariate Analysis

#### Sociodemographic Factors

Compared to those who had not had a prior COVID-19 test, those who had been tested were more likely to be living in San Diego County (versus Tijuana) and were less likely to identify as Hispanic, Latinx or Mexican. COVID-19 testing was positively associated with having completed high school or its equivalent and being homeless in the last six months (Tables 1 and 2).

**Table 2:** Factors associated with SARS-CoV-2 testing in Tijuana and San Diego.

| **Baseline Characteristics** | **Univariate OR**<br>**(95% CI)** |
| --- | --- |
| ***Sociodemographic Factors*** |  |
| Male<sup>P</sup> | 0.91 ( 0.61,1.36 ) |
| Age<sup>P</sup> | 1.00 ( 0.99,1.02 ) |
| Hispanic/Latinx/Mexican | 0.25 ( 0.17,0.37 ) |
| Speaks English | 5.06 ( 2.99,8.58 ) |
| Born in the US | 6.57 ( 4.33,9.97 ) |
| Primary residence in San Diego | 5.64 ( 3.62,8.79 ) |
| Homeless* | 2.07 ( 1.45,2.96 ) |
| Completed high school or its equivalent | 5.64 ( 3.62,8.79 ) |
| Monthly income <500 USD | 0.47 ( 0.33,0.68 ) |
| ***Behavioral Factors*** |  |
| # of hours spent on the street* | 1.04 ( 1.02,1.07 ) |
| Engaged in sex work*<sup>P</sup> | 0.79 ( 0.46,1.36 ) |
| ***Substance Use Factors*** |  |
| Smoked/snorted/inhaled/vaped methamphetamine* | 2.05 ( 1.39,3.03 ) |
| Smoked/snorted/inhaled cocaine* | 3.29 ( 1.94,5.56 ) |
| Smoked/snorted/inhaled/vaped fentanyl* | 4.38 ( 2.83,6.78 ) |
| Smoked/snorted/inhaled/vaped heroin* | 2.31 ( 1.58,3.40 ) |
| Injected methamphetamine* | 2.04 ( 1.42,2.91 ) |
| Injected fentanyl* | 2.43 ( 1.60,3.68 ) |
| Injected heroin*<sup>P</sup> | 0.80 ( 0.48,1.35 ) |
| Years of injection drug use<sup>P</sup> | 1.00 ( 0.99,1.02 ) |
| #Times injected drugs per day | 0.87 ( 0.78,0.98 ) |
| ***Mental Health and Stigma Factors*** |  |
| GAD-7 anxiety scale | 1.04 ( 1.01,1.07 ) |
| Anticipated Stigma (worry about experiencing increased verbal/physical harassment during COVID) | 1.63 ( 1.21,2.19 ) |
| Enacted Stigma (experienced increased verbal/physical harassment during COVID) | 1.72 ( 1.18,2.49 ) |
| ***Attitudinal Factors*** |  |
| On a scale of 1 to 10, how worried are you of getting COVID-19 (or getting it again) | 0.94 ( 0.88,0.99 ) |
| Does NOT think the virus that causes COVID-19 can be easily spread from one person to another<sup>Y3P</sup> | 0.87 ( 0.55,1.36 ) |
| Does NOT think that many thousands of people have died from COVID-19<sup>Y3P</sup> | 0.67 ( 0.38,1.19 ) |
| Thinks that most people already have immunity to COVID-19<sup>Y3P</sup> | 0.89 ( 0.60,1.31 ) |
| Thinks that you can tell someone has COVID-19 by looking at them<sup>Y3P</sup> | 1.05 ( 0.67,1.65 ) |
| Thinks that there are effective treatments for COVID-19 that can cure most people<sup>Y3</sup> | 0.66 ( 0.43,1.00 ) |
| Thinks that having COVID-19 is about as dangerous as having the flu<sup>Y3P</sup> | 1.31 ( 0.90,1.92 ) |
| ***Health-Related Factors*** |  |
| Ever had a flu vaccine<sup>Y2</sup> | 2.71 ( 1.85,3.97 ) |
| Tested HIV+ | 0.53 ( 0.25,1.12 ) |
| Tested HCV+ | 1.43 ( 1.00,2.05 ) |
| Has at least one chronic condition (excluding seasonal allergies and acne/skin problems) | 2.48 ( 1.73,3.57 ) |
| # of chronic conditions (excluding seasonal allergies and acne/skin problems) | 1.40 ( 1.19,1.65 ) |
| ***COVID-19-Related Factors*** |  |
| Income worse since COVID began<sup>Y1</sup> | 0.59 ( 0.41,0.86 ) |
| Low or very low food security since COVID began | 0.70 ( 0.45,1.08 ) |
| Exposed to someone with COVID-19 | 4.03 ( 1.97,8.25 ) |
| Knows someone who died of COVID-19<sup>Y5</sup> | 1.41 ( 0.95,2.08 ) |
| Reported being vaccinated for COVID-19 | 1.66 ( 1.01,2.75 ) |
| Tested SARS-CoV-2 seropositive<sup>Y6P</sup> | 1.05 ( 0.83,1.33 ) |
| ***COVID-19 Protective Behaviors*** |  |
| Social Distancing | 3.68 ( 2.52,5.39 ) |
| Wore face mask | 1.69 ( 1.05,2.73 ) |
| Most important source of COVID-19-related information: Friends<sup>Y4</sup> | 0.42 ( 0.28,0.63 ) |
| Most important source of COVID-19-related information: Doctors/health professionals<sup>Y4</sup> | 5.20 ( 2.50,10.8 ) |
| Most important source of COVID-19-related information: Social media<sup>Y4</sup> | 2.59 ( 1.52,4.43 ) |
| ***Touchpoint Opportunities for COVID-19 Testing*** |  |
| Incarcerated* | 3.19 ( 1.84,5.53 ) |
| Slept in a shelter/welfare residence* | 3.73(1.96, 7.10) |
| Overdose* | 1.88 ( 1.18,3.00 ) |
| Tested for HIV or HCV post-COVID | 2.10 ( 1.46,3.02 ) |
| Has been enrolled in a drug treatment program* | 3.56 ( 1.97,6.44 ) |
| Has been enrolled in a methadone or buprenorphine program* | 3.67 ( 1.85,7.25 ) |
| Attended a syringe service program* | 3.13 ( 1.07,9.16 ) |

\*Past 6 months; Missing values: <sup>Y1</sup> n=8, <sup>Y2</sup> n=41; <sup>Y3</sup> n=39; <sup>Y4</sup> n=50; <sup>Y5</sup> n=37; <sup>Y6</sup> n=12; <sup>P</sup>P-value>0.10 (all others ≤0.10)

#### Behavioral and Substance Use Factors

Behaviors significantly associated with higher odds of COVID-19 testing included non-injection use of fentanyl or injecting methamphetamine in the last six months and spending more time on the street.

#### Attitudinal Factors

Having a higher perceived threat of COVID-19 was associated with higher odds of COVID-19 testing. Endorsing most statements reflecting COVID-19 misinformation were not significantly associated with a lower odds of COVID-19 testing with the exception of believing that the coronavirus was created by the Chinese government as a biological weapon.

#### Mental Health and Stigma Factors

Increased anxiety reflected by higher GAD-7 scores was associated with higher odds of COVID-19 testing. Additionally, both anticipated stigma scores (i.e., worry of experiencing increased verbal or physical harassment during COVID-19 pandemic) and enacted stigma scores (e.g., feeling verbally or physically harassed since COVID-19 began) were associated with higher odds of COVID-19 testing.

#### Health-related Factors

Having diabetes, at least one chronic condition, ever having had a flu vaccine and testing HCV or HIV seropositive were significantly associated with COVID-19 testing.

#### COVID-19 related Factors

Using facemasks, practicing social distancing, having received at least one COVID-19 vaccine dose, and having been exposed to someone with COVID-19 were significantly associated with COVID-19 testing. Having primarily obtaining their COVID-19 information from social media or health providers was significantly associated with higher odds of COVID-19 testing, whereas obtaining most of their COVID-19 information from friends was inversely associated with COVID-19 testing.

#### Touchpoints for COVID-19 Testing

Having been incarcerated, overdosed, slept in a shelter, receiving SUD treatment or visiting a SSP program in the last six months were significantly associated with COVID-19 testing.

#### Factors Independently Associated with COVID-19 Testing

In our final multivariate model controlling for time, living in San Diego County (versus Tijuana), having been incarcerated or enrolled in a SUD treatment program in the last six months were independently associated with COVID-19 testing (Table 3). Having at least one chronic condition, receiving at least one COVID-19 vaccine dose, having been homeless or using fentanyl by means other than injection in the last six months were also independently associated with having had a COVID-19 test. Having been tested for HIV or HCV since the COVID-19 epidemic began remained marginally associated with COVID-19 testing.

**Table 3.**
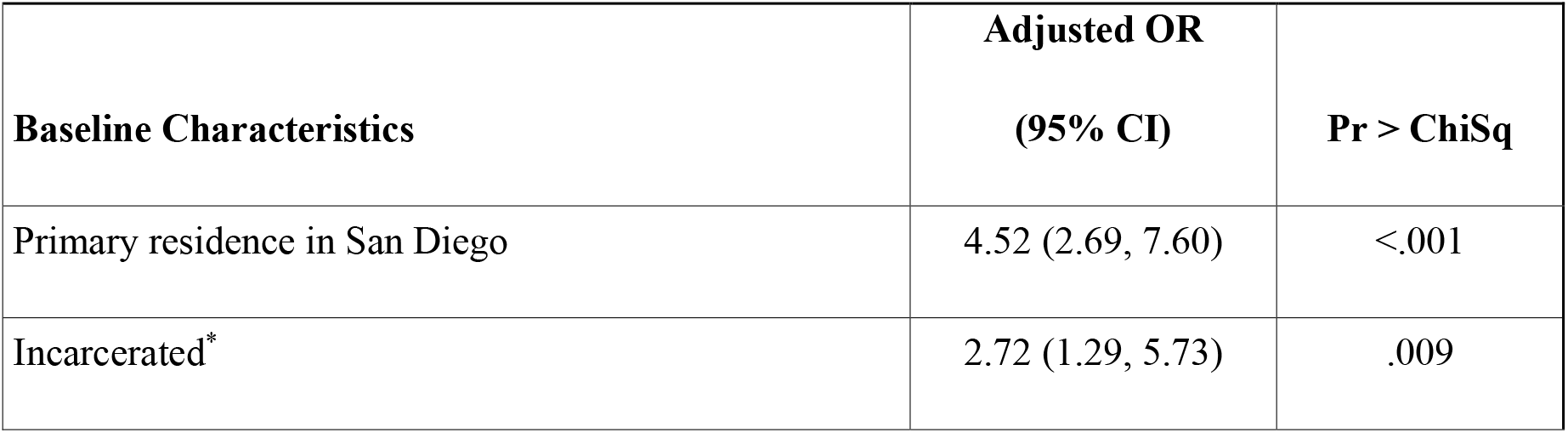
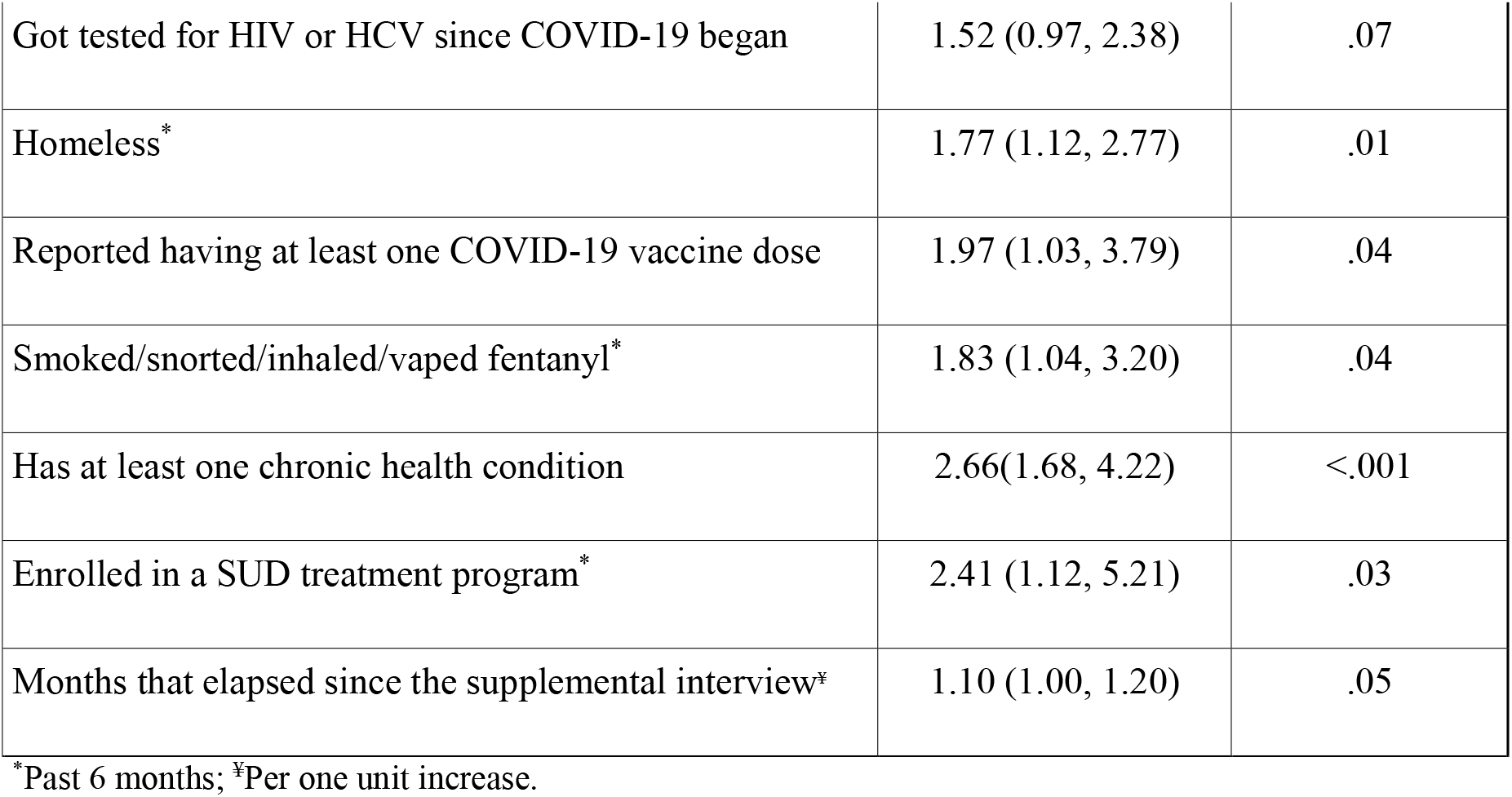
Factors Independently Associated with COVID-19 Testing among People who Inject Drugs in San Diego, CA and Tijuana, Mexico.

## Discussion

We identified several factors that were independently associated with COVID-19 testing among PWID in the Mexico-US border region, as well as numerous touchpoints where COVID-19 testing could have been offered. Although SARS-CoV-2 prevalence among PWID in San Diego County and Tijuana is higher than that of the general population in either city, [17] less than one third of our sample had ever been tested for COVID-19. Of concern, over two thirds of participants who tested SARS-CoV-2 seropositive in our study had not had a COVID-19 test and half reported at least one missed opportunity for testing. Our findings are consistent with a study of current and former substance users in Baltimore, Maryland, which found that only 13% had received a COVID-19 test in the first quarter of the pandemic. [15] Similarly, in a study of PWID in England and Northern Ireland conducted in 2020, only 22% ever had a COVID-19 test. [36] These findings have implications for improving service delivery for this vulnerable population, as well as broader efforts to reduce SARS-CoV-2 transmission and morbidity and mortality in the community.

An encouraging finding was that PWID who reported receiving SUD treatment were more likely to have been tested for COVID-19. This is supported by data from a recent study of 265 clients receiving residential SUD treatment in Southern California, among whom 74% had received a COVID-19 test. [24] Although it is not clear whether or not these individuals had received testing at the program itself or whether SUD treatment was a marker for health-seeking behaviors, some participants did report having had a COVID-19 test at SUD treatment clinics in our study. SUD treatment programs could serve as an ideal venue for providing COVID-19 testing as well as vaccines and education to dispel myths about COVID-19 testing and vaccination. However, during the pandemic, some SUD treatment programs were suspended or only offered take-home or telemedicine services, [37] potentially reducing opportunities for the provision of other services.

We also found that recent incarceration was associated with more than a two-fold higher odds of having had a COVID-19 test. Indeed, one in ten individuals who had a prior COVID-19 test reported that it occurred in a jail, prison or detention center. In many of these cases, COVID-19 testing may have been mandatory, since COVID-19 outbreaks have been reported in correctional facilities in California and elsewhere [38] and incarceration was independently associated with SARS-CoV-2 seropositivity among PWID in the same study sample. [17] These outbreaks prompted mass SARS-CoV-2 testing in some jurisdictions. [23] A study conducted in 2020 among the U.S. Federal Bureau of Prisons found that half of the prison populace had been subjected to COVID-19 testing. [39]

As expected, PWID who were already in contact with the healthcare system were more likely to have had a COVID-19 test. Those who had been tested for HIV or HCV since the COVID-19 epidemic began, and those who had received at least one COVID-19 vaccine dose were significantly more likely to have had a COVID-19 test. Furthermore, those who had at least one chronic condition were more likely to receive COVID-19 testing, which is noteworthy since individuals with co-morbidities are at greater risk of developing serious COVID-19 complications. [13]

Our findings that homelessness and non-injection use of fentanyl were both independently associated with having a COVID-19 test was surprising. However, this could be explained by concerted efforts in both San Diego and Tijuana to provide outreach to people experiencing homelessness during the COVID-19 pandemic, for example, through health fairs, mobile testing units and temporary housing. Similarly, if those using fentanyl had greater addiction severity, these individuals may have been more likely to come into contact with community-based health providers who offered COVID-19 testing. These interpretations are speculative and deserve greater attention. Kral and colleagues have documented marked transitions from injection of black tar heroin to non-injection use of fentanyl in San Francisco [40] and this sub-group of substance users may be more attuned to their health.

Contrary to expectation, we did not find that Latinx ethnicity was associated with a lower odds of COVID-19 testing; however, ethnicity was highly correlated with place of residence. We did not find that COVID-19 disinformation, misinformation or stigma were significantly associated with a reduced odds of COVID-19 testing after controlling for other factors, perhaps because these measures were not specific to COVID-19 testing. Future studies may benefit from the incorporation of tailored COVID-19 testing disinformation, misinformation, and stigma measures informed by qualitative research to more fully assess the influence of these factors on COVID-19 testing behaviors.

The lack of affordable and accessible rapid COVID-19 testing has been a major shortfall of the public health response to the pandemic in the United States and Mexico, [9, 41, 42] and elsewhere. Considering that over half of our participants earned less than $500 USD per month and the prevalence of homelessness was high, it is unreasonable to expect that this population would have access to financial or transportation resources (or sufficient access to the Internet) to be able to purchase expensive at-home test kits or make and attend appointments for COVID-19 testing. To reduce high rates of morbidity and mortality due to COVID-19 and ongoing SARS-CoV-2 transmission, it is critical that infections among PWID and other vulnerable populations are detected early, especially given their high rates of SARS-CoV-2 infection. [17] Efforts to expand free rapid testing for PWID at venues they already access and trust are especially needed, given recent evidence suggesting that people with SUD may be more prone to breakthrough infections following vaccination due to their high prevalence of co-morbidities. [43]

### Limitations

This study was limited by the cross-sectional nature of the analysis, which precludes causal inferences. Although this was a binational study, sampling was non-random and results may not generalize to other samples of PWID. Our survey relied on self-report and may have been subject to socially desirable responding or recall challenges. We could not differentiate between circumstances in which COVID-19 testing was mandatory (e.g., in prison, jails, detention centers, and some homeless shelters) versus situations in which testing was voluntary and sought out by participants. These contextual factors are important to explore in future studies. Similarly, we did not ask participants if they were required to pay for COVID-19 testing. Noting that the availability and cost of COVID-19 testing may have changed over time, we controlled for time in our analysis.

Statistical power may have limited our ability to detect some associations. For example, we did not find that attending SSPs was independently associated with COVID testing, which could be related to the low number of participants who had recently accessed SSPs. During the COVID-19 epidemic, many U.S. SSPs reduced their services or hours of operation, [25] or reduced provision of other health services. [44] In California, free rapid COVID-19 testing only became available through mobile SSPs after data collection for this study ended.

## Conclusions

Despite the high prevalence of SARS-CoV-2 among PWID in San Diego County and Tijuana, over two thirds of PWID had never had a COVID-19 test prior to testing SARS-CoV-2 seropositive in our study, and half reported encounters with at least one touchpoint where COVID-19 testing could have been offered. PWID were more likely to have been tested for COVID-19 if they had received health care access for other health conditions or had HIV or HCV testing, or if they had recently received SUD treatment or been incarcerated. Given the overall low level of COVID-19 testing and numerous missed opportunities where testing could be offered, there is an urgent need to improve access to free rapid COVID-19 tests in venues trusted and routinely accessed by PWID. Such initiatives may also improve uptake of COVID-19 vaccines.

## Data Availability

All data produced in present study are available upon reasonable request to the authors.

## Acknowledgements

The authors gratefully acknowledge the La Frontera study team and participants in San Diego and Tijuana and staff at Genalyte and Fluxergy for assistance interpreting laboratory results, laboratory staff at the Center for AIDS Research and Sharon Park for assistance with manuscript preparation.

## Declaration of Interests

The authors report no conflicts of interest.

## Data Deposition Information

For all data requests, please contact Steffanie Strathdee, Ph.D. at

## Notes

**Funding:** This work was supported by the National Institute on Drug Abuse (NIDA) at the National Institutes of Health (NIH) (R01DA049644-S1, K01DA043412, T32DA023356 and RADxUP, R01 DA049644-02S2). Additional support was provided by the National Institute of Allergy and Infectious Diseases (P30 AI036214) and the Fogarty International Center (R21DA051951).

### Competing Interest Statement

The authors have declared no competing interest.

### Funding Statement

This work was supported by the National Institute on Drug Abuse (NIDA) at the National Institutes of Health (NIH) (R01DA049644-S1, K01DA043412, T32DA023356 and RADxUP, R01 DA049644-02S2). Additional support was provided by the National Institute of Allergy and Infectious Diseases (P30 AI036214) and the Fogarty International Center (R21DA051951).

### Author Declarations

Office of IRB Administration at the University of California San Diego (Project# 191390)

